# Lesions Causing Aphantasia are Connected to the Fusiform Imagery Node

**DOI:** 10.1101/2025.05.23.25328072

**Authors:** Julian Kutsche, Calvin Howard, Alberto Castro Palacin, William Drew, Matthias Michel, Alexander L. Cohen, Michael D. Fox, Isaiah Kletenik

## Abstract

The absence of visual mental imagery, called aphantasia, occurs congenitally in up to 3% of the general population, but the brain regions responsible for aphantasia remain uncertain. Rare cases of acquired aphantasia caused by brain lesions may lend insight into the neuroanatomy responsible for this condition, and the neural substrate of visual mental imagery itself.

We performed a systematic literature review to identify cases of lesion-induced aphantasia and traced the lesion locations onto a common brain atlas. These locations were compared to control lesions causing other neuropsychiatric symptoms (*n =* 887). First, we tested for intersection between lesion locations and an a priori region of interest termed the *fusiform imagery node*, active during visual mental imagery tasks. Second, we tested for connectivity between lesion locations and this region of interest, leveraging resting-state functional connectivity from a large cohort of healthy subjects (*n* = 1000). Finally, we performed a data-driven analysis assessing whole-brain lesion connectivity that was sensitive (100 % overlap) and specific (family-wise error *p* < 0.05) for aphantasia.

We identified 12 cases of lesion-induced aphantasia, only 5 of which intersected the *fusiform imagery node*. However, 100% of these lesion locations were functionally connected to the *fusiform imagery node*. Connectivity to this region was both sensitive (100% overlap) and specific (family-wise error *p*<0.05) for aphantasia in a data-driven whole-brain analysis.

Lesions causing acquired aphantasia occur in multiple different brain regions but are all functionally connected to the left *fusiform imagery node*. This study provides causal support for the importance of this brain region in visual mental imagery.

## 1. Introduction

Visual mental imagery, the ability to volitionally form perceptual representations without corresponding external sensory stimuli, is a unique cognitive function that allows for reliving past events, solving problems and envisioning the future (Pearson et al., 2015). Recent work has highlighted a broad range of visual mental imagery abilities across the human population identifying some extremes. The absence of voluntary visual mental imagery, called ‘aphantasia,’ has been recognized to occur congenitally in about 1–3 % of the general population (Dance et al., 2022; Zeman, 2024).

Current fMRI research in healthy participants has identified a focal region in the left ventral visual pathway thought to be specialized for visual mental imagery termed the *fusiform imagery node* (Spagna et al., 2021). Some studies have shown this region to be active during mental imagery tasks and altered in people with congenital aphantasia (Liu & Bartolomeo, 2025; Liu et al., 2025; Zeman, 2024). While these studies demonstrate correlative evidence for this region, causal inference is lacking (Hajhajate et al., 2022). If the fusiform imagery node is specific for visual mental imagery, then damage or disconnection of this region would abolish this function.

By studying rare cases of lesion-induced aphantasia, we can: 1) assess evidence for brain regions causally relevant for conscious visual mental imagery, and 2) provide guidance on which brain locations are likely to cause loss of visual mental imagery when injured by stroke or traumatic brain injury.

Since lesions causing aphantasia are in different locations across the brain, we apply the technique of *lesion network mapping* (Cohen et al., 2019; Kletenik et al., 2022; Kletenik, Gaudet, et al., 2023), which allows mapping the connectivity of lesion locations based on a large connectivity atlas. Here, we provide the first group-level network approach to mapping acquired aphantasia and identify regions specific for visual mental imagery.

## 2. Methods

We conducted a systematic literature review on PubMed following PRISMA guidelines using search terms related to stroke or other brain injury and aphantasia or visual mental imagery (see PRISMA checklist and Supplementary Materials). The literature review was conducted using the following search terms on articles published between 1980 and 2025: “(“stroke” OR “brain ischemia” OR “cerebral ischemia” OR “apoplexy” OR “intracranial hemorrhage” OR “intracranial haemorrhage” OR “cerebral hemorrhage” OR “brain hemorrhage” OR “lesion” OR “brain lesion” OR “acquired” OR “loss of”) AND (“aphantasia” OR “aphantasic” OR “mind’s eye” OR “mental visualization” OR “mental imagery” OR “visual imagery”)”. Findings were checked manually and with an automated search tool (Howard, 2025).

Our inclusion criteria were: 1) a loss of visual mental imagery, 2) after a brain injury, and 3) a published image of sufficient quality for mapping. We found 254 papers for review which identified 9 cases in 8 studies, and 3 additional cases were found through related citations (**see Supplementary Figure 1, Figure 1, and Table 1**). Brain lesions were traced by JK and reviewed for accuracy by two experienced neurologists (IK, CH).

**Figure 1:**
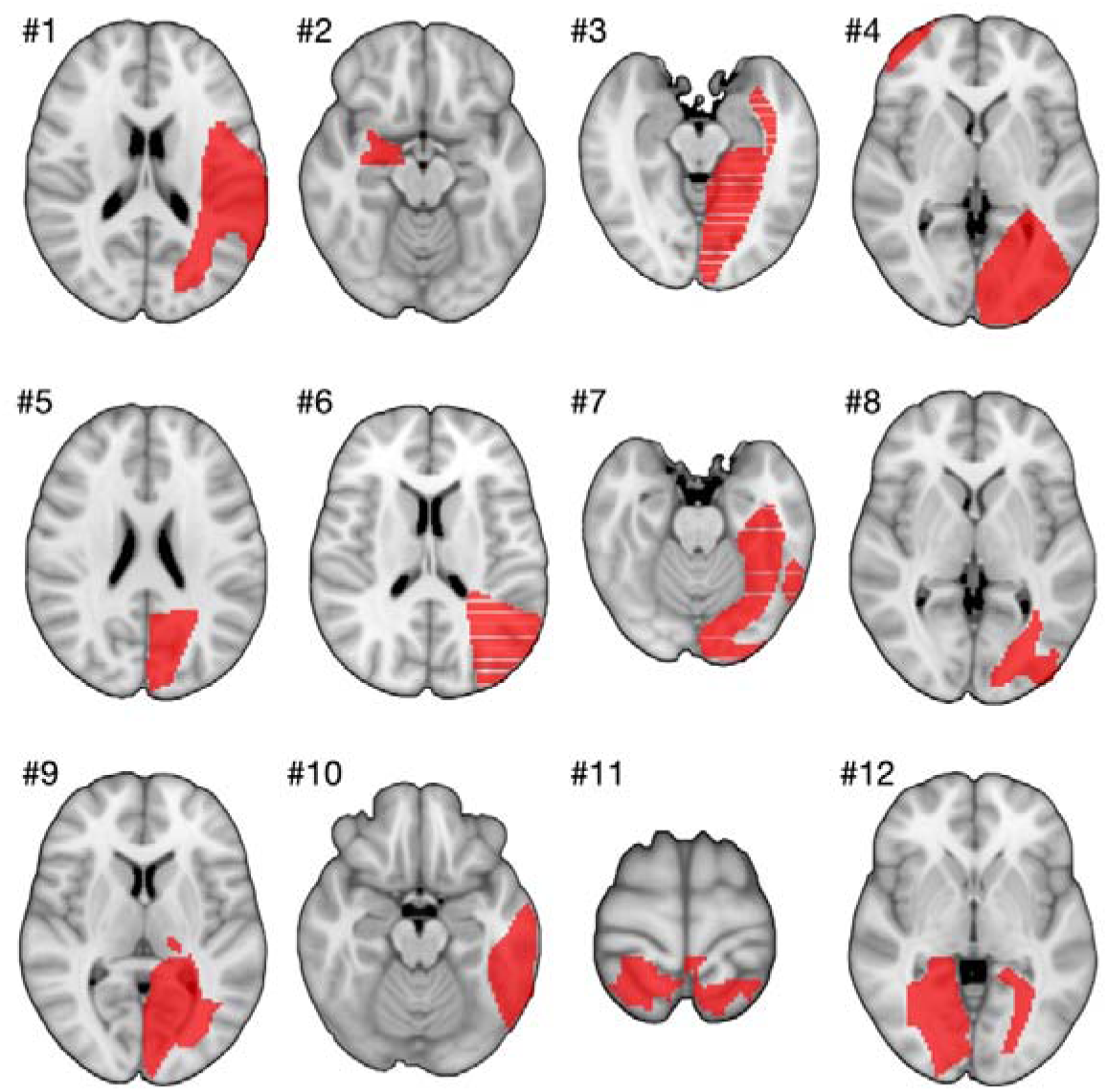
Lesion-induced aphantasia: Mapped locations (red) of 12 cases of lesion-induced aphantasia. Most lesions were left-lateralized.

**Table 1.**
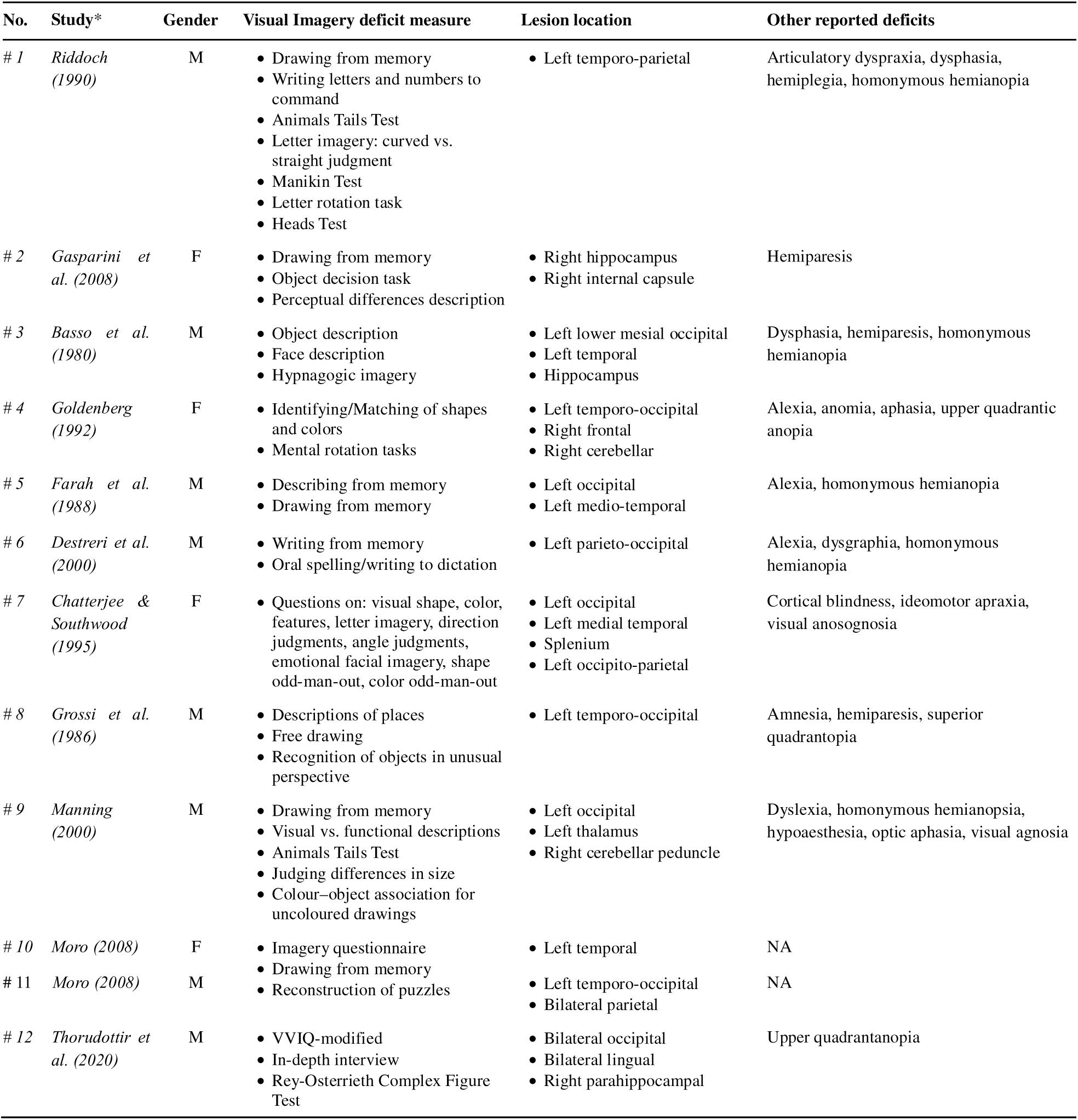
Table of aphantasia case demographic, anatomic and visual imagery information.

### 2.1 Hypothesis-driven approach: Lesion intersection and functional connectivity to fusiform image node

To test our hypothesis that disruption of the fusiform imagery node causes loss of visual mental imagery, we analyzed both the lesion intersection and lesion connectivity to this location. We defined the fusiform imagery node as the conjunction between the significant clusters from a recent ALE meta-analysis (Spagna et al., 2021) and cytoarchitectonic area FG4 from JuBrain atlas (Lorenz et al., 2017) consistent with recent work (Hajhajate et al., 2022; Spagna et al., 2021; Spagna et al., 2024). First, we assessed the difference in lesion intersection with the fusiform imagery node between aphantasia lesions and controls, which included all symptom-causing lesions analyzed in our lab (*n* = 887) and repeated the analysis with 100 randomly selected control lesions (to ensure results were not driven by group imbalance) using logistic regression. Group membership (aphantasia versus controls) served as the binary dependent variable while lesion intersection with FIN was the independent variable; lesion volume was added as a co-variate in a secondary analysis. Then, we calculated the ROI-to-ROI correlation between the fusiform imagery node and aphantasia lesions and the same group of controls controls using a large-scale functional connectome (*n* = 1000 subjects) to obtain lesion-wise average *r*-values. We transformed the *r*-values into *fz*-values using Fisher *z*-transformation and performed a two-tailed 2-sample *t*-test of average *fz*-values to this region for aphantasia lesions versus controls consistent with prior work (Kutsche et al., 2025). Additionally, we repeated this ROI-to-ROI correlation with an independently derived FIN location (Liu et al., 2025).

### 2.2 Data-driven approach: Whole-brain lesion network mapping

Functional connectivity was calculated using a previously validated method termed *lesion network mapping* (Cohen et al., 2019; Kletenik et al., 2022; Kletenik, Gaudet, et al., 2023), where for each patient’s lesion or set of lesions, average connectivity at each voxel in the brain is calculated using a publicly available 1000-subject resting-state functional connectome **(Figure 2A**).

**Figure 2:**
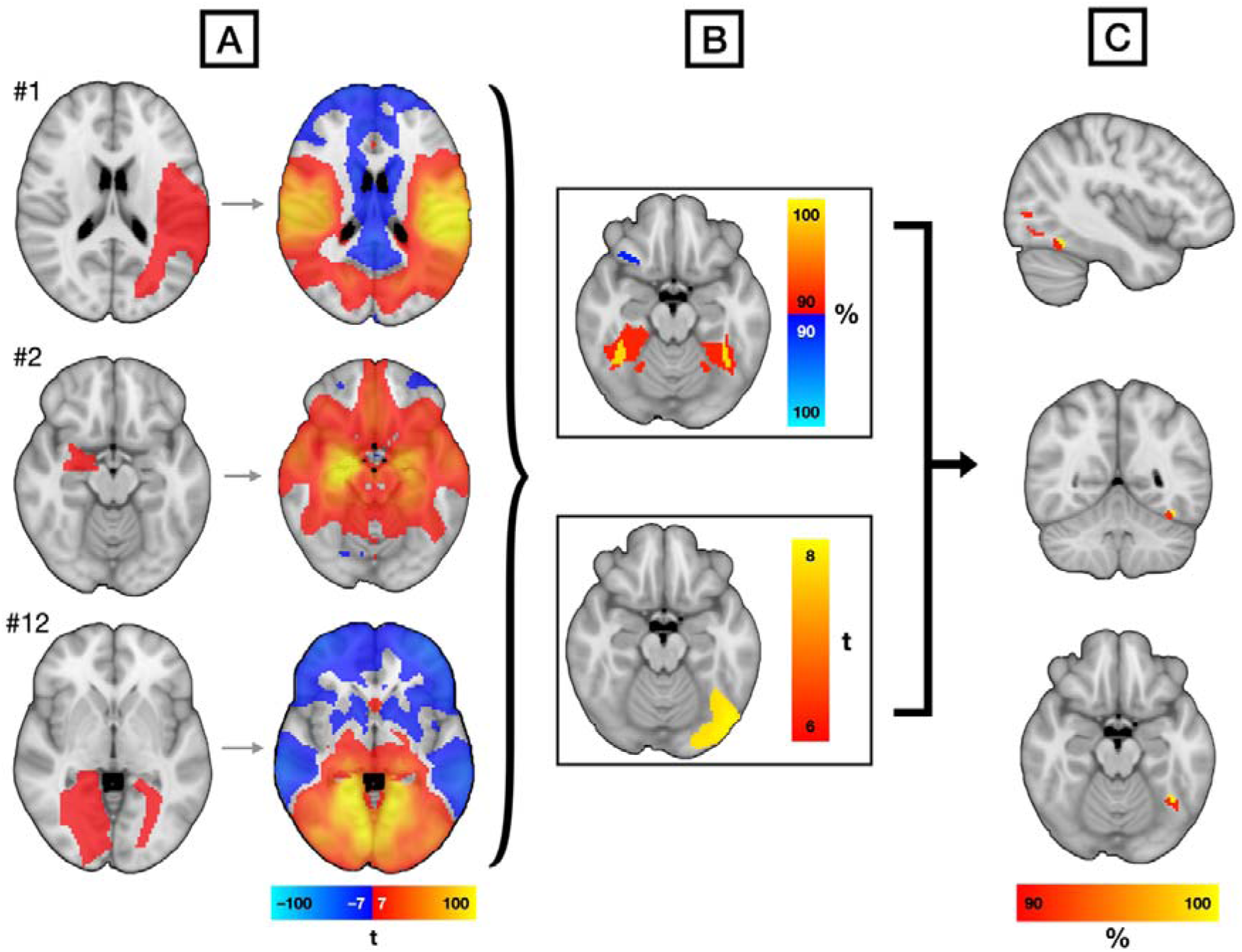
Methodology for functional lesion network mapping of aphantasia. (**A**) Three representative lesions causing loss of visual mental imagery (left) show injury to multiple different brain regions. Functional connectivity of each lesion location (right) was calculated using a large resting-state functional connectivity atlas. (**B**) Sensitivity analysis (top) and specificity analysis (bottom) show that aphantasia lesions are connected to the fusiform gyrus. (**C**) Conjunction of the sensitivity and specificity analyses identifies a focal region in the left fusiform gyrus that defines an aphantasia-causing network.

### 2.3 Sensitivity and Specificity

Sensitivity was assessed by calculating a *network overlap map*, where each lesion’s connectivity *t-*map is thresholded (*t* ≥ 7), binarized, and overlaid to show overlap connectivity (**Figure 2B, top**); this threshold is consistent with prior work and is not biased by control selection. (Cohen et al., 2019) To test for specificity, network maps for aphantasia were compared via whole-brain voxel-wise two-tailed 2-sample *t*-test employing Permutation Analysis of Linear Models (Winkler et al., 2014) to all symptom-causing lesions analyzed in our lab (*n* = 887), controlling for group membership, and with a threshold for significance of family-wise error (FWE) *p* < 0.05 (**Figure 2B, bottom**) (Kutsche et al., 2025). The conjunction of these two maps defines a sensitive and specific network hub for aphantasia (**Figure 2C**).

### 2.4 Secondary Analyses

To ensure that our aphantasia results in visual selective regions (such as the FIN) was not due to the diversity of control case symptoms, we repeated our whole brain voxel-wise two sample *t*-test comparing aphantasia lesions just to visual symptom controls (lesions causing cortical blindness, visual anosognosia, prosopagnosia, visual hallucinations and blindsight) from our control dataset. Note that some of these visual symptoms could have unrecognized co-occurring aphantasia so they are not the ideal test.

The relationship between aphantasia and prosopagnosia is particularly debated (Dance et al., 2023; Grüter et al., 2009; Monzel et al., 2023). Because we found that 23 % of prosopagnosia-causing lesions (Cohen et al., 2019) also intersect the FIN, and both aphantasia and prosopagnosia have reported localization to the region of the fusiform face area (FFA) with differing data on lateralization (prosopagnosia often to the right and aphantasia to the left) we compared these two groups directly. To do this, we performed a voxel-wise, two-tailed 2-sample *t*-test using PALM between the 12 aphantasia lesion network maps and 44 lesion network maps for prosopagnosia (Cohen et al., 2019) masked to a region of interest of the bilateral fusiform face area as defined by a validated, human visual category-selective atlas (Rosenke et al., 2021) to assess differences in lateralization between these conditions.

### 2.5 Comparison to prior work

We then calculated the functional connectivity of our sensitive and specific aphantasia location, using the same connectome, to produce an aphantasia network. We compared this aphantasia network to: 1) the functional connections of the independently derived *a priori* fusiform imagery node using spatial correlation and 2) the deactivation foci from an independent functional neuroimaging study of a person with acquired aphantasia performing visual mental imagery tasks (Zeman et al., 2010). The hypothesis is that positive connectivity of lesions causing acquired aphantasia from our network would align with deactivation coordinates from this fMRI (Kutsche et al., 2025) from a subject with acquired aphantasia.

### 2.6 Bayesian Analysis

Although frequentist testing can identify which networks are most consistently associated with a lesion-induced symptom, null findings in this framework do not distinguish between insufficient evidence and genuine non-involvement. To address this limitation, we also incorporated Bayesian hypothesis testing, which allows formal comparison of models that include or exclude candidate regions. This approach enables us to quantify whether our data provide affirmative evidence for the absence of involvement of specific regions, such as the primary visual cortex, fusiform cortex or frontal lobes, in a way that frequentist mapping approaches cannot.

In this Bayesian analysis, we used the Bayesian Spatial Generalised Linear Mixed Model (BSGLMM) (https://www.nisox.org/Software/BSGLMM/) (Ge et al., 2014). Consistent with prior work and the thresholds we used in our frequentist sensitivity analysis, network maps were thresholded at *t* > 7 and binarized as inputs. BSGLMM estimates voxel-wise posterior mean probabilities that reflect the probability, given the data and the spatial Bayesian model, that each voxel shows an effect associated with the aphantasia-causing network group. In this analysis, we focused on the aphantasia-causing network posterior mean probability map where the range is from 0 to 1. To facilitate comparison with the conjunction of the sensitivity and specificity results, we applied a threshold of posterior mean probability > 0.95, yielding a spatial map that is conceptually similar in interpretability to the family-wise-error-thresholded output. Although a posterior mean probability > 0.95 is not statistically equivalent to a family-wise-error-corrected *p*-value < 0.05, both approaches identify voxels with strong evidence for an effect. Voxels surviving this threshold therefore represent locations where the Bayesian model provides high confidence that the lesion network associated with aphantasia involves these regions. We therefore used the 0.95 probability threshold to enable a qualitative comparison with the FWE-corrected *p*-values results. Clusters, coordinates and anatomic regions surviving this threshold were identified using MRIcroGL and the Automated Anatomical Labeling (AAL) atlas with the standard minimum cluster size of 32mm^3^. Finally, we examined the overlap of suprathreshold voxels between the two methods to assess the degree of convergence between frequentist and Bayesian inference. Voxels identified by both statistical frameworks represent the regions with the highest probability of being involved in the neural mechanisms underlying aphantasia.

### 2.7 Structural connectivity approach

Prior work has left uncertainty regarding white matter connections specific to visual mental imagery and aphantasia (Milton et al., 2021). Hence, we complemented our functional connectivity analysis with a structural connectivity approach. Using BCBToolkit (Foulon et al., 2018), we derived structural connectivity maps for each of the 12 aphantasia lesions and 887 control lesions using a large diffusion MRI (dMRI) structural connectome (*n* = 178) (Vu et al., 2015). To closely approximate our functional connectivity analysis, we employed both a sensitivity and specificity analysis. For the structural sensitivity analysis, connectivity maps were binarized at the toolkit’s default threshold (> 50 %) and overlaid to obtain a structural network overlap. Then, for the specificity analysis we employed these structural network maps unthresholded in a whole-brain voxel-wise 2-sample *t-*test comparing the aphantasia cases to the same controls (Kletenik, Cohen, et al., 2023; Kletenik et al., 2025). Note that this 7T dMRI connectome is in 1 mm space which was used for the structural connectivity analysis.

## 3. Results

We identified 12 cases of lesion-induced aphantasia (mean age 59.25 year [SD 17.8], 4 female) which were primarily due to stroke (**Figure 1 and Table 1**) (Basso et al., 1980; Chatterjee & Southwood, 1995; Destreri et al., 2000; Farah et al., 1988; Gasparini et al., 2008; Goldenberg, 1992; Grossi et al., 1986; Manning, 2000; Moro et al., 2008; Riddoch, 1990; Thorudottir et al., 2020). Lesions were mostly left lateralized but otherwise occurred in many different regions including frontal (4/12), parietal (9/12), temporal (11/12), and occipital lobes (12/12) and subcortical locations (8/12) (**Figure 1**).

### 3.1 Hypothesis-driven approach: lesions causing aphantasia are connected to the fusiform imagery node

Only 42 % of aphantasia lesions directly intersected the fusiform imagery node, though significantly more than control lesions of which 4 % intersected this region (β = 2.95, *p* < 0.000001, OR = 19.09; when correcting for lesion volume: β = 3.66, *p* < 0.000001, OR = 38.82; or compared to 100 randomly selected control lesions: β = 2.61, *p* < 0.001, OR = 13.6). In contrast, 100 % of the aphantasia lesions were functionally connected to the fusiform imagery node in our region of interest analysis. Aphantasia lesions also showed significant connectivity to the fusiform imagery node (Spagna 2021), compared to control lesions (*t*_11.104_ = 3.7442, *p* < 0.005, Cohen’s *d* = 1.81, **Figure 3A**) and results were similar (*t_897_* = 5.4741, *p* < 0.005, Cohen’s *d* = 1.59) using an independently derived fusiform imagery node (Liu et al., 2025). In addition, aphantasia lesions had a stronger connectivity to the fusiform imagery node than any of the other neuropsychiatric symptoms; some visual syndromes, especially prosopagnosia, also showed connectivity to this region (**Supplementary Figure 2**), consistent with prior work (Dance et al., 2023; Zeman et al., 2020).

**Figure 3.**
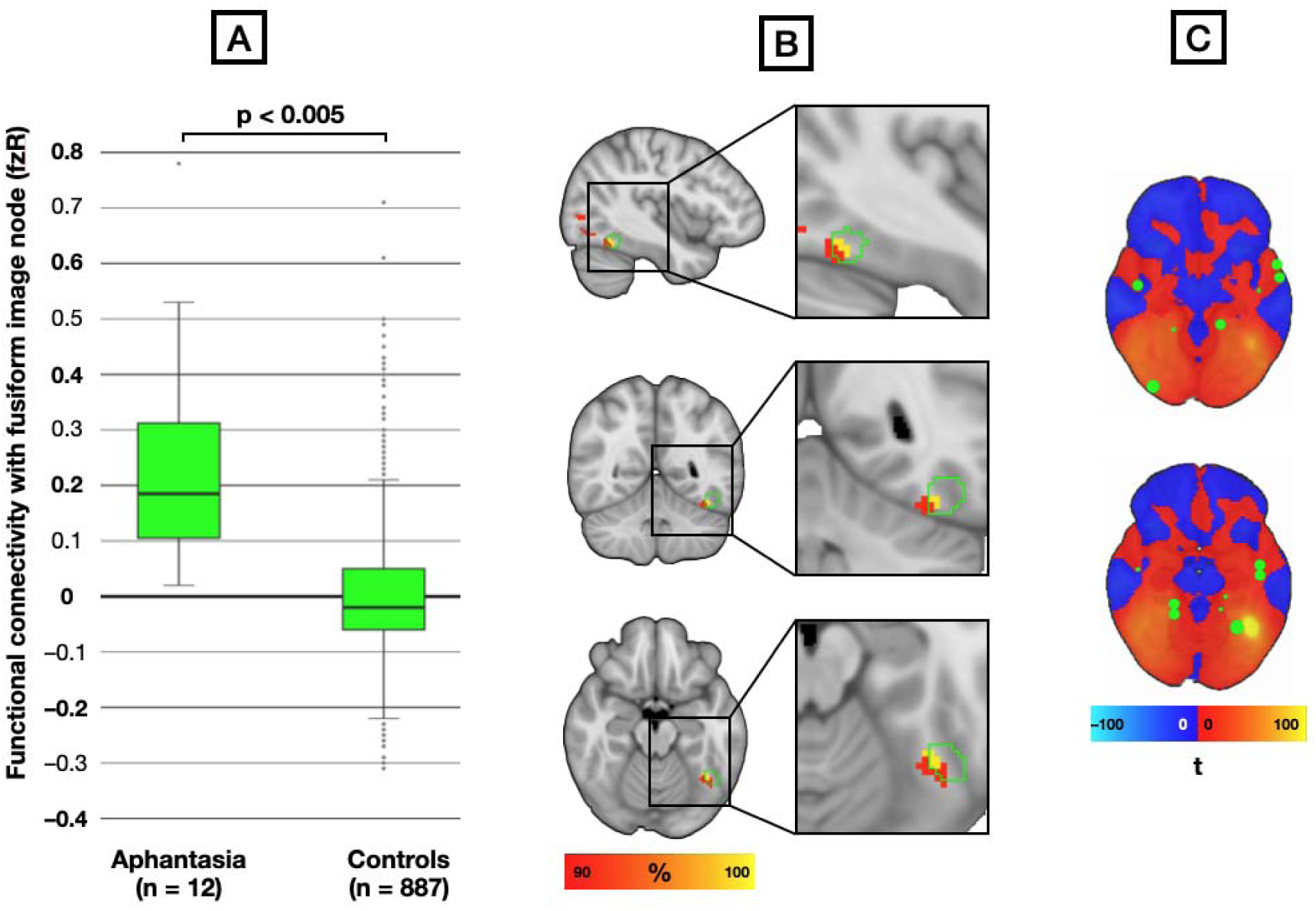
Lesions causing aphantasia are all functionally connected to the left fusiform imagery node. (**A**) Comparison of fusiform imagery node connectivity to lesions causing aphantasia versus 887 control lesions causing diverse neuropsychiatric symptoms. Lesions causing aphantasia show a significant functional connectivity correlation to the fusiform imagery node compared to 887 control lesions causing diverse neuropsychiatric symptoms (*t* = 3.7442, p<0.005). (**B**) The whole-brain, data-driven, aphantasia network demonstrates a region that all aphantasia-causing lesions (FWE p < 0.05, 100% of lesion network overlap) are functionally connected to (yellow) which is primarily within an independently derived region recently termed the *fusiform imagery node* (outlined in green, per Spagna et al.(Spagna et al., 2021)). (**C**) Locations of decreased activity (green) during an fMRI visual mental imagery task from a person with acquired aphantasia (Zeman et al., 2010) intersect with positive connections from lesions that cause aphantasia.

### 3.2 Data-driven functional approach: The fusiform imagery node was sensitive and specific for aphantasia

We then applied a whole-brain approach to determine locations sensitive and specific for aphantasia without *a priori* assumptions. On functional sensitivity analysis, 100 % of aphantasia lesions were functionally connected to the fusiform gyri (**Figure 2B, top**). For our specificity analysis, we compared aphantasia lesions to controls with 24 diverse neuropsychiatric symptoms employing whole-brain voxel-wise 2-sample *t*-test; aphantasia lesions were connected to the left temporo-occipital fusiform cortex (FWE *p* < 0.05, **Figure 2B, bottom**). The conjunction of these networks identified a region both sensitive and specific for aphantasia in the left inferior fusiform gyrus (peak MNI coordinate: *x* = –38, *y* = –56, *z* = – 18, **Figure 2C**), of which 85 % of voxels were within the area of the *fusiform imagery node* (**Figure 3B**) (Spagna et al., 2021). Functional connectivity with this location defined an aphantasia network.

Our aphantasia lesion network showed strong spatial correlation with the functional connectivity of the ALE meta-analysis (Spagna et al., 2021) fusiform peak from studies of normal subjects performing visual mental imagery tasks (*r* = 0.86; see **Supplementary Figure 3**). Finally, our aphantasia lesion network aligned with the deactivation foci from a person with acquired aphantasia (21/24 foci; **Figure 3C**) (Zeman et al., 2010).

### 3.3 Relationship between the aphantasia network and other visual symptoms

In a secondary analysis, the aphantasia network peak location was similarly within the fusiform imagery node (peak MNI coordinate: *x* = –50, *y* = –56, *z* = –16) when compared to just visual symptom controls (uncorrected *p* < 0.01). In addition, contrasting aphantasia and the prosopagnosia network maps on a voxel-wise 2-sample *t*-test, masked to the fusiform face area, demonstrated that aphantasia lesion connectivity lateralized to the left fusiform face area while prosopagnosia lesion connectivity lateralized to the right (uncorrected *p* < 0.05, **Supplementary Figure 4**).

### 3.4 Bayesian analysis

Bayesian analysis identified a focal set of voxels with strong posterior evidence for involvement in aphantasia. Applying a posterior mean probability threshold of > 0.95 yielded a spatial map of voxels primarily within the bilateral fusiform and inferior temporal gyri (**Supplementary Figure 5A, Supplementary Table 1**). No significant clusters were observed within the frontal lobes or the calcarine cortex (V1). The areas of convergence between Bayesian and frequentist approaches fell in the left fusiform gyrus nearly entirely within FIN (Supplementary Figure 5B).

### 3.5 Data-driven structural connectivity approach

We then repeated our whole-brain approach employing a structural connectome. The structural sensitivity map identified an inferior left-lateralized white matter network. Because a threshold of 100 % did not leave any conjunction between the sensitivity and specificity maps, a threshold of 91.7 % (11/12 cases) was employed. For the structural specificity map we compared the aphantasia lesions to the same controls with 24 diverse neuropsychiatric symptoms employing whole-brain voxel-wise 2-sample *t*-test. Aphantasia lesions were connected to a structural network in the left temporo-occipital white matter (FWE-*p* < 0.05, **Figure 4A**). The conjunction of our structural sensitivity and structural specificity networks was a white matter connection superior and posterior to the fusiform imagery node primarily within the left inferior longitudinal fasciculus (peak MNI coordinate: *x* = –34, *y* = –71, *z* = –5; **Figure 4C**).

**Figure 4.**
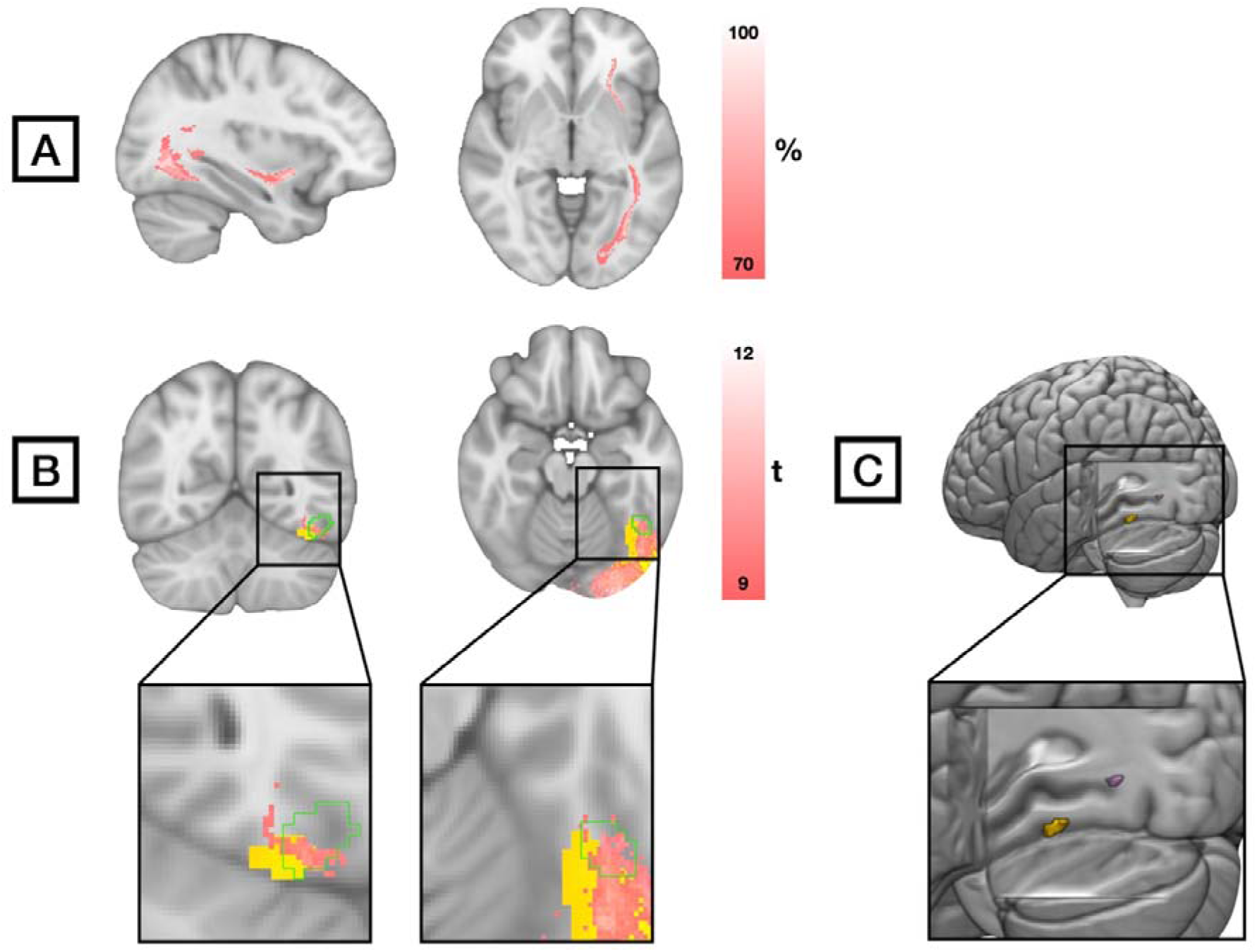
Structural lesion network mapping of aphantasia. (**A**) Structural sensitivity analysis of aphantasia which shows the structural network overlap of lesions causing aphantasia based on percent of cases. (**B**) Specificity of significant structural connectivity locations (pink) of aphantasia lesions compared to 887 control lesions (FWE-*p* < 0.05). Also shown for comparison are the specificity results from the functional connectivity analysis of aphantasia versus controls (yellow, Figure 2B**, bottom**) and the FIN (Spagna 2021, green). (**C**) Topographical relation of the conjunction maps of sensitive and specific structural (pink) and functional (yellow) connectivity peaks in a 3D brain.

We cannot directly compare the structural and functional network topography using spatial correlation, as the two results were derived from different connectomes and datatypes (Kletenik, Cohen, et al., 2023); however, the topography of our aphantasia structural specificity network was qualitatively similar to our aphantasia functional specificity network (**Figure 4B**).

## Discussion

Lesions causing loss of visual mental imagery were all functionally connected to the left fusiform imagery node. This location defined a brain network that was sensitive and specific for aphantasia. Consistent with prior work, our results lateralized to the left hemisphere and our sensitive and specific results were focused in the ventral visual association cortex (Bridge et al., 2012; D’Esposito et al., 1997; Hahamy et al., 2021; Spagna et al., 2021; Zeman, 2024). Our findings were robust across different methodologies and add disease-associated, causal data to prior work on the importance of the fusiform imagery node for visual mental imagery tasks in healthy participants (Spagna et al., 2021).

Why is the location of the fusiform imagery node so essential for voluntary visual mental imagery? This region may be a specialized location in the left fusiform gyrus for processing imagined visual representations (D’Esposito et al., 1997). Alternatively, the fusiform imagery node may be uniquely situated to connect regions involved in semantic processing (especially the left anterior temporal lobe) with memory regions (including medial temporal lobe) to ventral visual cortex thereby allowing a person to voluntarily evoke a specific semantic concept, access stored memories of it and employ them to form a visual mental representation (Spagna et al., 2021). Prior research has shown that deactivation in the left fusiform gyrus is associated with suppression of visual memories (Gagnepain et al., 2014) which may also explain why lesions connected to this region lead to loss of the ability to evoke visual mental images which rely on visual memory. Little is known about the structural connections associated with acquired aphantasia and our structural connectivity conjunction finding, adjacent to FIN within the left inferior longitudinal fasciculus, may emerge, with further study, as a key white matter connection for voluntary visual mental imagery.

The fusiform imagery node was not only sensitive and specific for aphantasia in our typical frequentist lesion network mapping analysis but remained a consistent finding in Bayesian hypothesis testing. Importantly, no significant clusters (in both our frequentist conjunction analysis and Bayesian analysis) were observed within the frontal lobes (Liu & Bartolomeo, 2025; Zeman, 2024) or primary visual cortex (Cabbai et al., 2024; Milton, 2024; Pearson, 2019) providing affirmative evidence against their specific involvement in acquired aphantasia. Why did we not identify activity in these regions? Our analysis is the first, group-level study of lesion-induced aphantasia, so the neural correlates of acquired aphantasia may be different than the correlates of visual mental imagery in normal participants or people with congenital aphantasia. Furthermore, while prefrontal activation (Zeman, 2024) may be part of a general process to initiate visual mental imagery tasks, connectivity to prefrontal regions may not be specific to lesion-induced aphantasia when compared to controls. In a similar vein, activity in the primary visual cortex has been linked to the vividness of visual mental imagery but it has not been sufficient to explain subjective imagery experience (Cabbai et al., 2024); V1 may then have a supportive role in visual mental imagery but injury to V1 would not abolish conscious visual mental imagery (Bridge et al. 2012). Our findings of the fusiform imagery node as both sensitive and specific to acquired aphantasia, with evidence against the involvement of other debated regions, including the lateral prefrontal cortex (Liu & Bartolomeo, 2025; Zeman, 2024) and V1 (Cabbai et al., 2024; Milton, 2024; Pearson, 2019), contributes new data to an ongoing debate on the neural underpinnings of visual mental imagery and aphantasia.

Our findings are relevant to neural correlates of consciousness research more broadly as lesions are able to disrupt conscious visual imagery experience. The exact relationship between the experience of visual mental imagery and objective performance on visual cognitive tasks is a key area of study that can help frame this discussion. Individuals with congenital aphantasia can perform quite normally on visual cognitive tasks that would seem to require visual imagery (Liu & Bartolomeo, 2023; Pounder et al., 2022). In some cases, this performance might be explainable by non-conscious, imagery-like visual representations in early visual cortex (Weber et al., 2024), whereby early visual cortex activity occurs unconsciously and is not sufficient for the conscious experience of visual imagery (Michel et al., 2025). Our result is consistent with this perspective and indicates that activity in the FIN is necessary for the conscious experience of visual imagery. There could be two explanations for this finding. One possibility is that the FIN is both necessary and sufficient for the conscious experience of visual imagery - an explanation consistent with so-called “local” views of consciousness (Malach, 2021). Another possibility is that the FIN is a necessary part of a broader set of regions, including perhaps regions in prefrontal cortex (Michel, 2022), that together support the conscious experience of visual imagery - in line with views of consciousness such as global workspace theory (Mashour et al., 2020) and higher-order theories (Brown et al., 2019). These frontal regions though would not be specific to aphantasia compared to controls nor would injury to frontal lobes specifically abolish visual mental imagery as in our cases of acquired aphantasia. There may also be important differences in the neural correlates and visual performance abilities in patients with congenital versus acquired aphantasia which requires more study of this rare condition (Arcangeli & Bartolomeo, 2025; Bartolomeo et al., 2002).

## Limitations

Our analysis relied on cases from the medical literature. Because of this our study had a small number of aphantasia participants, most participants lacked full vividness scales or objective aphantasia measures and there were case-based differences in imagery terminology (Blomkvist & Marks, 2023). Despite this, our results were statistically robust in a whole-brain analysis with a conservative threshold, but our small sample size may have limited our significant results to peak regions. We also relied on limited 2-dimensional images;, however, prior lesion network mapping studies have shown that 2-dimensional slices can accurately approximate connectivity patterns of whole lesions (Boes et al., 2015; Kletenik, Gaudet, et al., 2023). Furthermore, while our results add important causal evidence to the role of the fusiform imagery node in visual mental imagery, functional connectivity changes in acquired aphantasia may be different than in the far more common congenital aphantasia (Arcangeli & Bartolomeo, 2025). More rigorous testing of visual mental imagery abilities in patients with stroke-related onset of aphantasia using validated measures is needed.

## Conclusion

Lesions causing aphantasia were all functionally connected to a region in the left ventral visual pathway, termed the *fusiform imagery node.* Our results provide causal evidence supporting the hypothesis that this region’s connections are key for voluntary visual mental imagery.

## Data Availability

Data produced are available online at

https://doi.org/10.7910/DVN/ILXIKS

https://doi.org/10.5281/ZENODO.15465733

https://neurovault.org/collections/ILPECICK/

https://www.humanconnectome.org/study/hcp-young-adult/document/1200-subjects-data-release

## Data availability

To compute functional connectivity, we used a publicly available resting-state functional connectome: https://doi.org/10.7910/DVN/ILXIKS and for structural connectivity we used a publicly available dMRI structural connectome: https://www.humanconnectome.org/study/hcp-young-adult/document/1200-subjects-data-release. Lesion and fMRI data used in this analysis are publicly available (previous ALE meta-analysis (Spagna et al., 2021) for fusiform imagery node mask, References for lesion studies). Statistical neuroimaging analyses were performed in Matlab, version R2024b (Mathworks Inc), SPSS 28.0.0.0 (IBM) and FSL (version 6.0.7.16). Other statistical analyses were run using R (version 4.4.3) using *psych* (Revelle 2025) and *lsr* (Navarro 2015) packages. Brain images were created using FSLeyes (version 1.13.0). Brain maps generated in our analyses can be publicly accessed on Neurovault: https://neurovault.org/collections/ILPECICK/

## Acknowledgments

We thank the many patients who participated in the included studies and to the researchers who publicly shared their results.

## Supplementary material

Supplementary material is available online.

## Ethical Standards

The study was approved by Mass General Brigham/ Partners Institutional Review Board Protocols 2015P001248, 2020P002987 and 2020P000737 and all participants provided written informed consent per individual journal requirements. This study has been approved by the appropriate ethics committee and has been performed in accordance with the ethical standards laid down in the 1964 Declaration of Helsinki and its later amendments.

