## Supplementary Tables and Figures for "Lesions Causing Aphantasia are Connected to the Fusiform Imagery Node"

Supplementary Materials

### **Supplementary Figure 1. PRISMA diagram for literature research on lesions causing aphantasia. Three additional cases were found in Spagna (2022).**

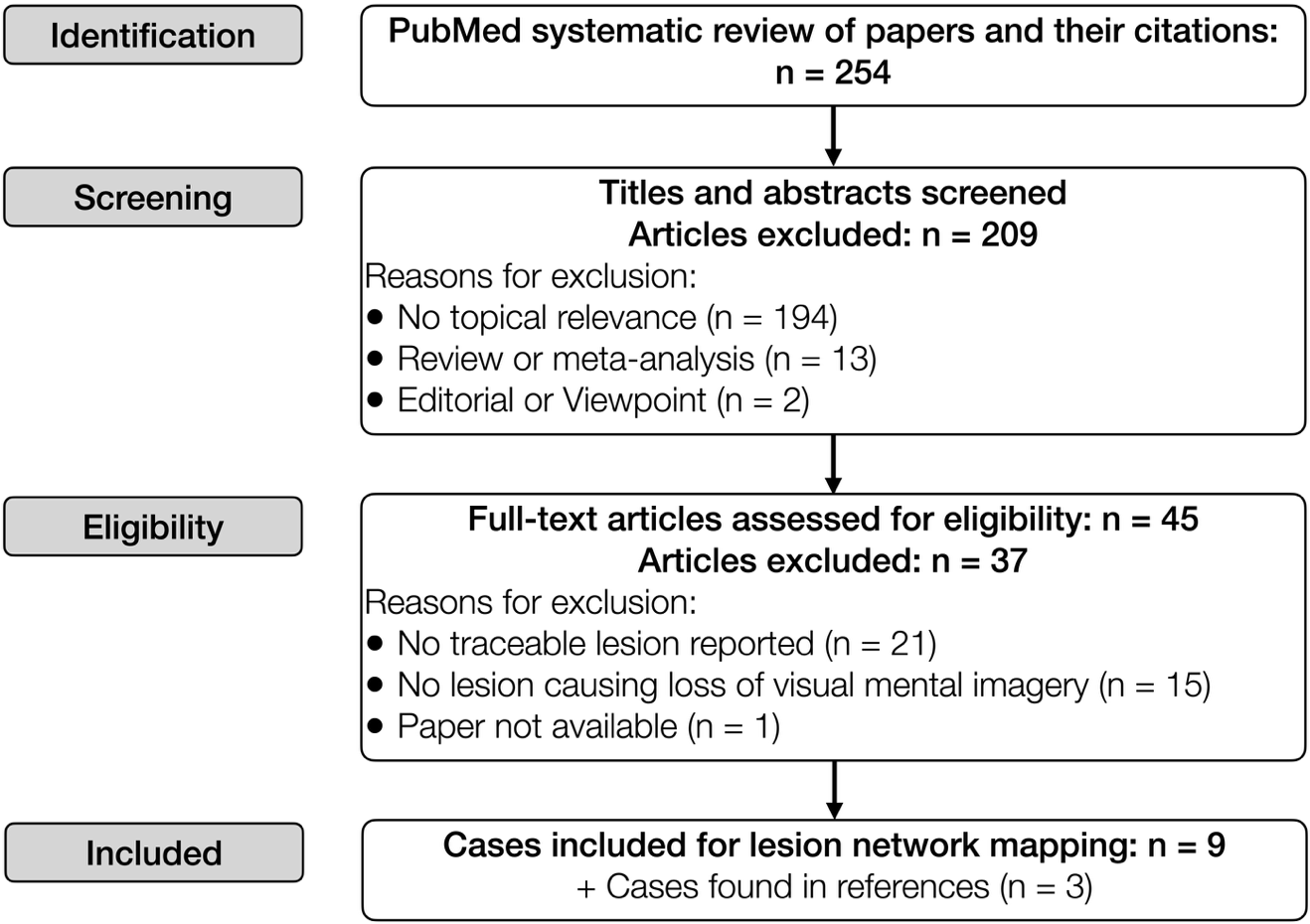

### **Supplementary Figure 2. Comparison of fusiform imagery node connectivity (per Spagna et al., 2021) to lesions causing aphantasia and 24 diverse neuropsychiatric symptoms.**

The fusiform imagery node shows the strongest functional connectivity correlation with lesions causing aphantasia followed by prosopagnosia and to a lesser extent other visual syndromes. Given the recent recognition of aphantasia and visual mental imagery scales, people with lesion-induced prosopagnosia and other visual syndromes may have unrecognized concurrent aphantasia.

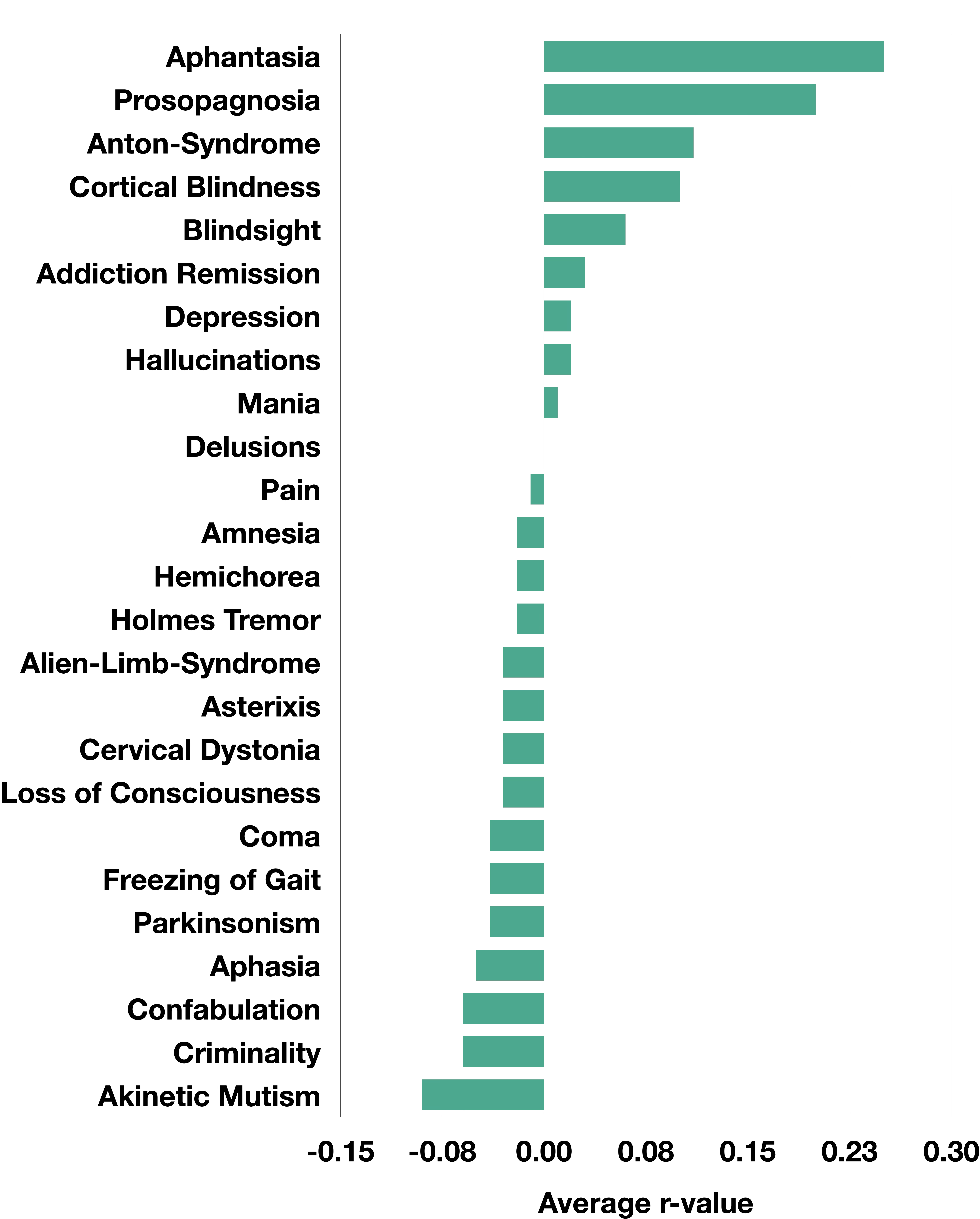

### **Supplementary Figure 3.** Topographical similarity of the functional connectivity of the fusiform imagery node (per Spagna et al., 2021) and the sensitive and specific aphantasia lesion location (*r* = 0.86).

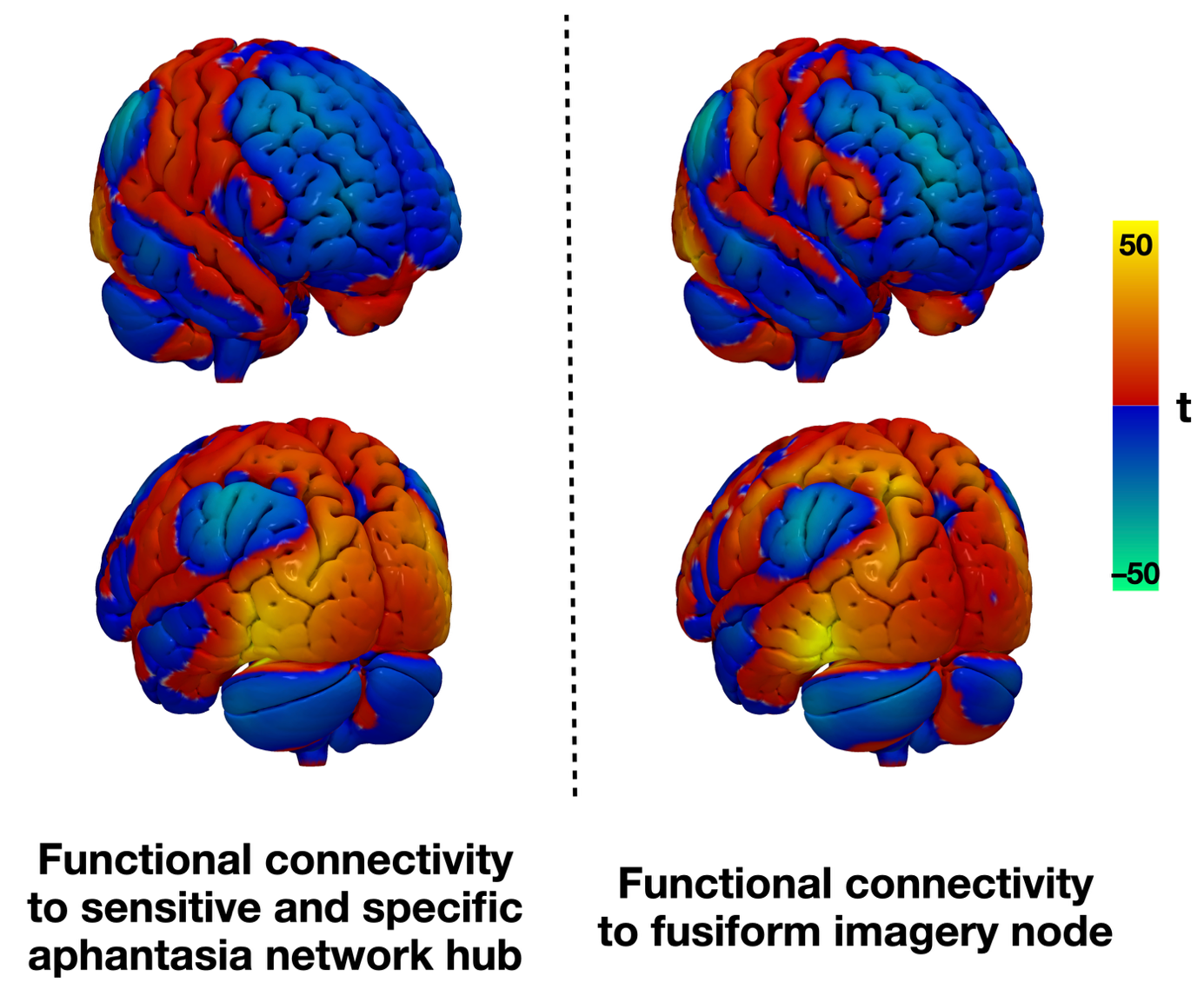

**Supplementary Figure 4. Comparison between aphantasia and other visual syndromes.**
(**A**) Voxel-wise comparison of aphantasia (warm colors) versus prosopagnosia lesion connectivity (Cohen et al., 2019), cool colors) masked to fusiform face area (white outline) (Rosenke et al., 2021). Lesions causing aphantasia show connections lateralized to the left FFA while prosopagnosia lesion connections lateralize to the right FFA. (**B**) Secondary, whole brain voxel-wise analysis of aphantasia causing lesions versus visual syndrome controls (*n* = 225), shows that aphantasia localizes to left fusiform even when compared just to visual symptoms with FIN (green outline) for comparison.

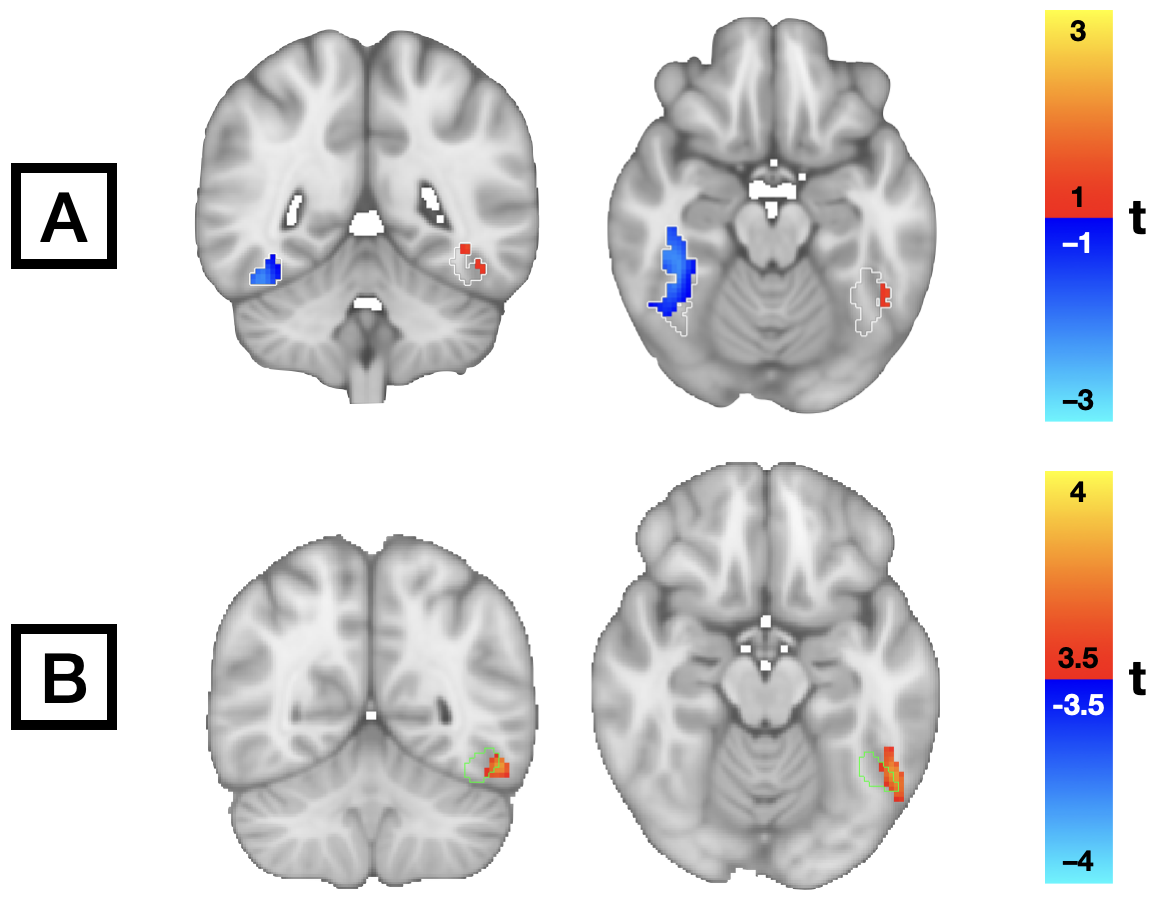

### **Supplementary Figure 5. Whole brain Bayesian analysis of aphantasia versus controls** (**A**) A whole-brain Bayesian analysis reveals a 95 % probability that voxels in temporo-occipital cortex, belong to the aphantasia group compared to the control group. None of the supra-threshold voxels fell within the frontal lobes (dark blue outline) or the primary visual cortex (red outline). All clusters were located within the bilateral fusiform (pink outline) and inferior temporal gyri (light blue outline) (**B**) A conjunction between the aphantasia network hub to which 100 % of aphantasia lesions are functionally connected and the Bayesian analysis (yellow) shows topographical alignment with the fusiform imagery node (green outline).

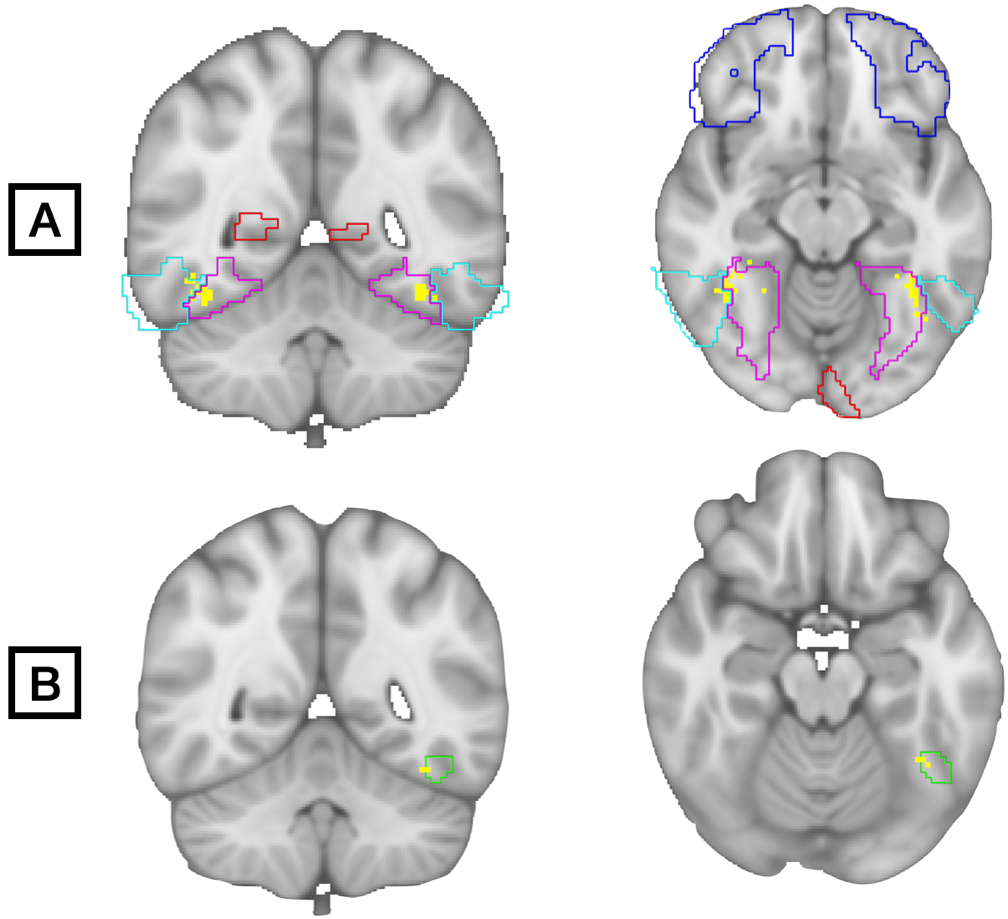

**Supplementary Table 1. Significant clusters from whole brain Bayesian analysis of aphantasia versus controls based on Automated Anatomical Labeling (AAL) atlas.**

| **Cluster size (mm^3^)** | **MNI coordinates** | **Structures** |
| --- | --- | --- |
| 714 | 41, –48, 17 | Right fusiform gyrus, Right inferior temporal gyrus |
| 598 | 50, –69, –6 | Right inferior temporal gyrus, Right inferior occipital gyrus |
| 307 | –37, –50, –16 | Left fusiform gyrus, Left inferior temporal gyrus |
| 61 | –36, –39, –21 | Left fusiform gyrus |
| 52 | –49, –71, 9 | Left middle occipital gyrus,  Left middle temporal gyrus |
